# Early Identification of Patients with Acute Gastrointestinal Bleeding using Electronic Health Record Phenotyping

**DOI:** 10.1101/2020.07.06.20136374

**Authors:** Dennis Shung, Cynthia Tsay, Loren Laine, Prem Thomas, Caitlin Partridge, Michael Simonov, Allen Hsiao, Andrew Taylor

## Abstract

**Background and Aim:** Guidelines recommend risk stratification scores in patients presenting with gastrointestinal bleeding (GIB), but such scores are uncommonly employed in practice. Automation and deployment of risk stratification scores in real time within electronic health records (EHRs) would overcome a major impediment. This requires an automated mechanism to accurately identify (“phenotype”) patients with GIB at the time of presentation. The goal is to identify patients with acute GIB by developing and evaluating EHR-based phenotyping algorithms for emergency department (ED) patients.

**Methods:** We specified criteria using structured data elements to create rules for identifying patients, and also developed a natural-language-processing (NLP)-based algorithm for automated phenotyping of patients, tested them with tenfold cross-validation (n=7144) and external validation (n=2988), and compared them with the standard method for encoding patient conditions in the EHR, Systematized Nomenclature of Medicine (SNOMED). The gold standard for GIB diagnosis was independent dual manual review of medical records. The primary outcome was positive predictive value (PPV).

**Results:** A decision rule using GIB-specific terms from ED triage and from ED review-of-systems assessment performed better than SNOMED on internal validation (PPV=91% [90%-93%] vs. 74% [71%-76%], P<0.001) and external validation (PPV=85% [84%-87%] vs. 69% [67%-71%], *P*<0.001). The NLP algorithm (external validation PPV=80% [79-82%]) was not superior to the structured-datafields decision rule.

**Conclusions:** An automated decision rule employing GIB-specific triage and review-of-systems terms can be used to trigger EHR-based deployment of risk stratification models to guide clinical decision-making in real time for patients with acute GIB presenting to the ED.

## Introduction

Acute gastrointestinal bleeding (GIB) is the most common gastrointestinal diagnosis requiring hospital admission in the United States.^1^ Guidelines for upper and lower GIB recommend risk stratification of patients, including the use of risk assessment scores.^2-4^ Although many risk stratification tools have been developed and validated, they are not commonly used in real-world clinical practice partly because providers must manually enter a variety of variables into the scoring system. Widespread electronic health record (her) adoption makes it possible to automatically deploy risk stratification scores within the clinical workflow for acute GIB; however, in order to embed risk stratification models into EHRs and deploy them in real time, patients must first be correctly identified. This process of accurately identifying patients, called phenotyping, is used for any study that seeks to reliably group patients with a specific diagnosis or condition, from surveillance studies to comparative effectiveness research. Phenotyping is typically the first step in developing and validating risk stratification models within EHRs.^5^ Such processes have been used to improve the accuracy of case definition of inflammatory bowel disease patients as Crohn’s disease or ulcerative colitis, as well as to facilitate clinical trial recruitment and deploy randomized controlled trials.^6, 7^

Unlike many conditions that require multiple elements of the record (laboratory testing, reported symptoms, and biometrics such as vital signs) for diagnosis, acute GIB is a condition that can be directly and clearly identified using a limited number of terms by patient report or provider evaluation. To our knowledge, however, no previous study has explored the early identification of patients with acute GIB with an EHR-based model or its implementation within standard EHR workflow.

Phenotypes utilize both structured and unstructured data and are typically used retrospectively after the clinical encounters have ended (e.g. ICD codes are a popular component of phenotypes). However, if the goal is to identify patients that would benefit from predictive models tailored for a particular condition, EHR phenotypes must use data elements generated during the visit. The Systematized Nomenclature of Medicine (SNOMED) is an international comprehensive clinical terminology that is the standard for encoding patient conditions in the EHR. Other approaches for phenotyping can be decision rules using specific data elements (e.g. triage diagnosis) or a machine learning (ML) approach to utilize unstructured clinical text through natural language processing (NLP). ML models use computational modeling to learn from data and their performance can improve with increasing amounts of data. NLP is a tool used to extract data from narrative text, and provides a comparison to decision rules. For prediction, however, a “screening” phenotype using data elements likely to be entered close to real time may be better than a tool that may be delayed (e.g. due to delays in notewriting).^8^

Our study aimed to accurately identify patients with acute GIB reported or witnessed in the emergency department, such that the identification of this phenotype can occur in real time in the EHR to subsequently launch predictive models for risk stratification. We chose to use the current standard for phenotyping patients, SNOMED, as the comparator, even though SNOMED does not provide real-time phenotyping in the emergency department.

## Methods

We began with creating a sensitive data mart, a patient dataset selected using specified criteria, to screen for all patients presenting with GIB from 2014 to 2017 in the Yale New Haven Health System electronic health record (Epic). Creation of a sensitive data mart allows exclusion of patients with no evidence of the phenotype and to adequately handle the volume, heterogeneity, and velocity of data.^9^ To create the data mart, we screened for patients with data that suggested the phenotype of overt gastrointestinal bleeding. In order to maximize the capture of relevant data, we defined the process by which patients were evaluated in the emergency department and common time periods for data entry. We also identified points at which a diagnosis would be entered in the electronic health record throughout the hospital stay: hospital problem list, encounter diagnosis, admission diagnosis, and hospital billing diagnosis.

There were 4 categories of screening criteria, which were selected based on existing identifiers in the EHR used to denote gastrointestinal bleeding (Figure 1).

**Figure 1:**
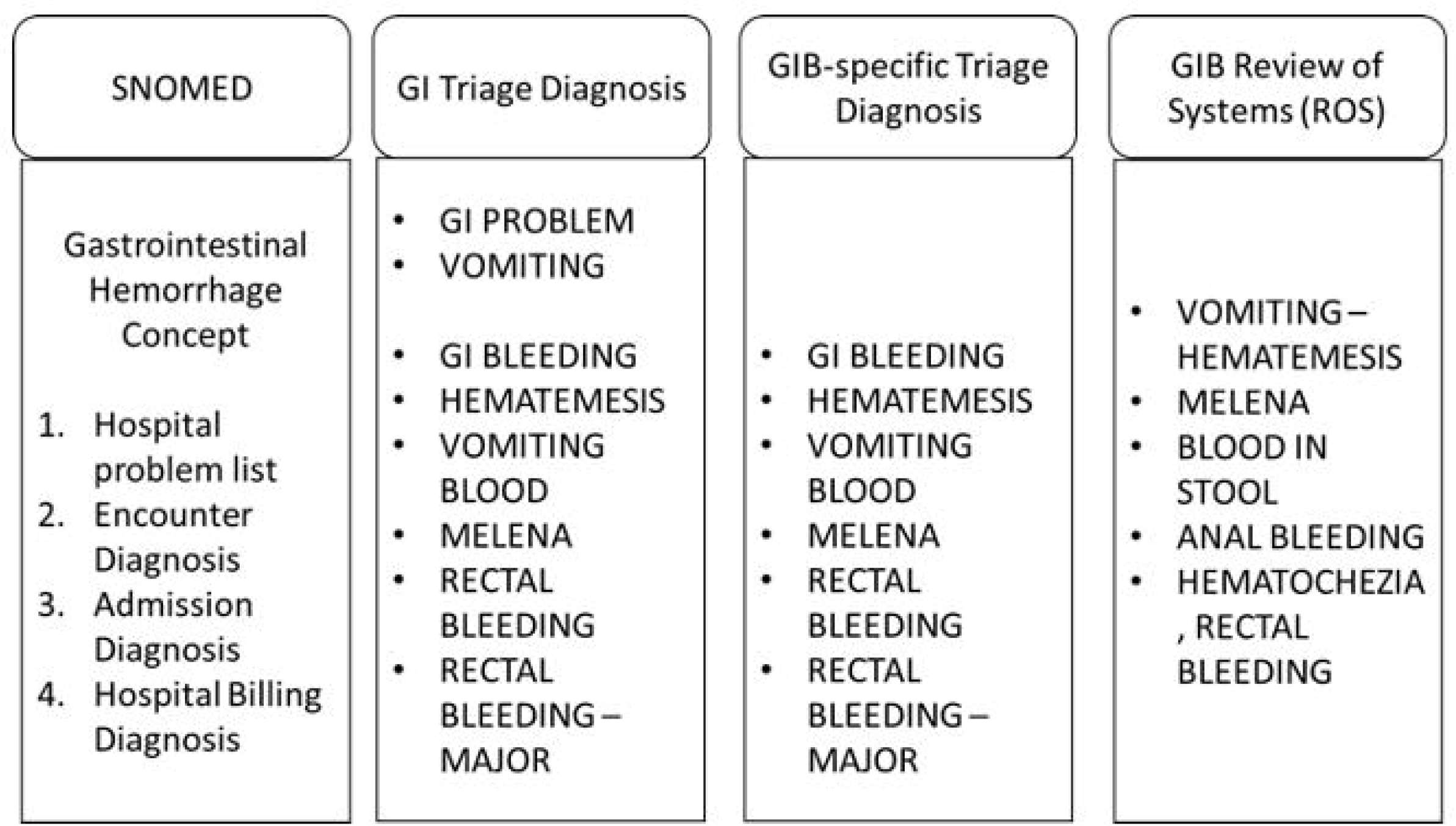
Screening criteria used to create data mart for acute gastrointestinal bleeding

### Development of EHR GIB Phenotypes

We developed rule-based algorithms and a machine-learning based algorithm using NLP and compared their performance to the SNOMED-only classification (Figure 2).

**Figure 2:**
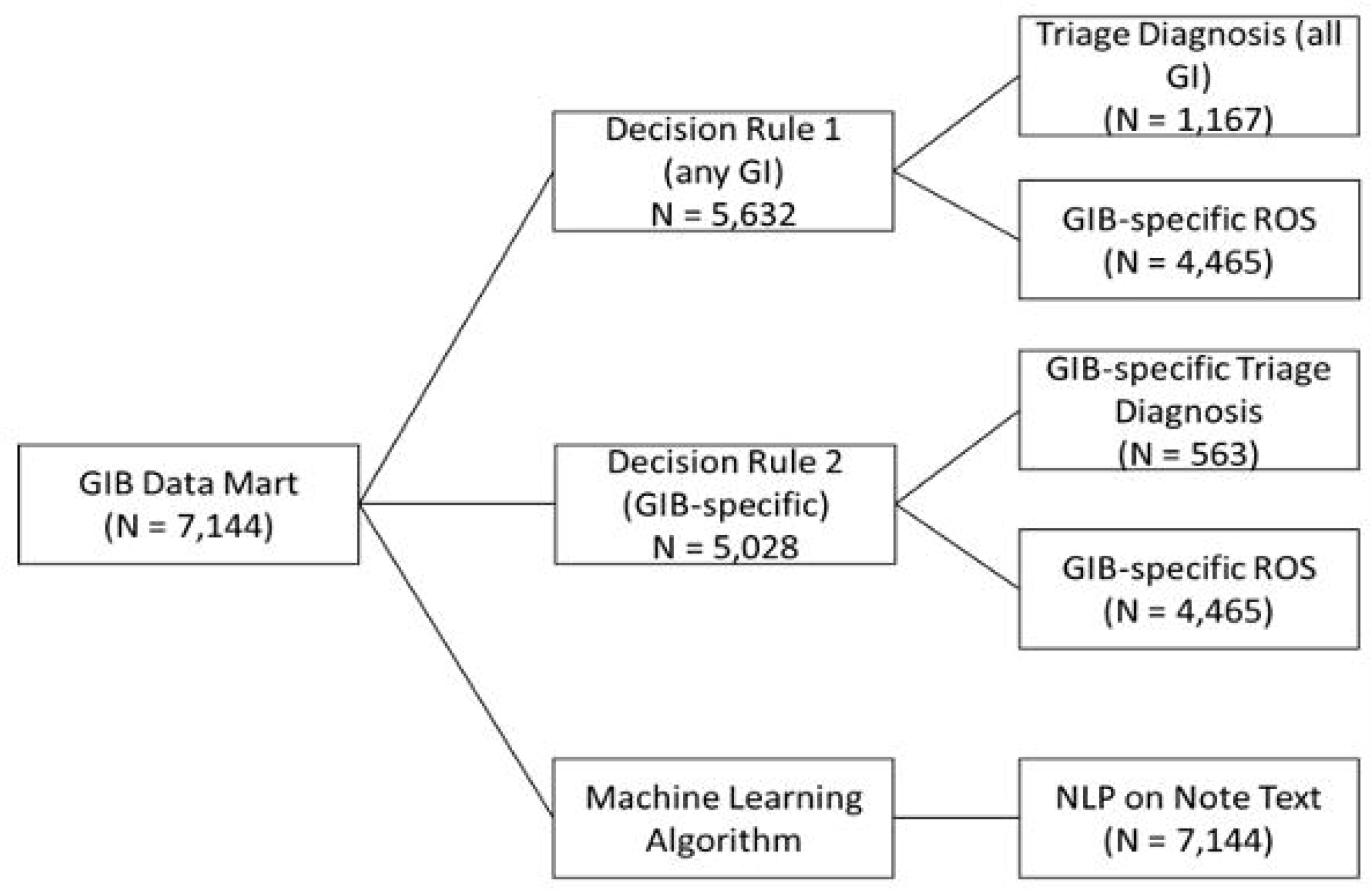
Rule-based algorithms and machine learning (Natural Language Processing) algorithm

To determine the specific data elements included in the rule-based algorithm, we analyzed the clinical workflow to identify two points where relevant diagnosis or symptom data was entered into the EHR. The first point of data entry was at triage, where a nurse selected presenting diagnoses from a drop down list of pre-specified diagnoses. We reviewed all triage diagnoses to identify any that were gastrointestinal diagnoses. The second data entry point was the review of systems section in the note template used for all ED patients, which contains elements referring to overt gastrointestinal bleeding. Both the triage diagnosis and review of systems provide structured data fields that can be used to identify patients as either 0 (not present) or 1 (present). We hypothesized that terms specific to gastrointestinal bleeding may have improved performance and therefore created two decision rules. Decision rule 1 was positive if the ROS field (e.g. hematemesis, blood in stool) was positive or if any GI triage term was positive. Decision rule 2 was positive if the ROS field was positive or GIB triage terms were positive.

The NLP approach includes preprocessing of the unstructured text in notes written by physician providers in the Emergency Department using ScispaCy, a Python software library used for advanced NLP that allows for breaking down text into smaller unique parts, by removing adverbs, pronouns, conjunctions, punctuation, participles and spaces.^10^ The text was then vectorized into bigrams and classified using a random forest classifier, a strategy that has been applied previously to radiology reports and other biomedical text data.^11,12^ We also performed a sensitivity analysis for hematemesis or melena and hematochezia since there is a distinction between upper gastrointestinal bleeding and lower gastrointestinal bleeding.

### Evaluation of Phenotype

To assess the performance of the different phenotypes, we performed manual note review to create a gold standard. Two clinical domain experts (DS and CT) reviewed the medical records of all patients and classified each patient as having the phenotype or not based on expert opinion and a prespecified structured evaluation (Table 3). We further categorized the acute bleeding by symptoms, either hematemesis/melena or hematochezia.

**Table 1:**
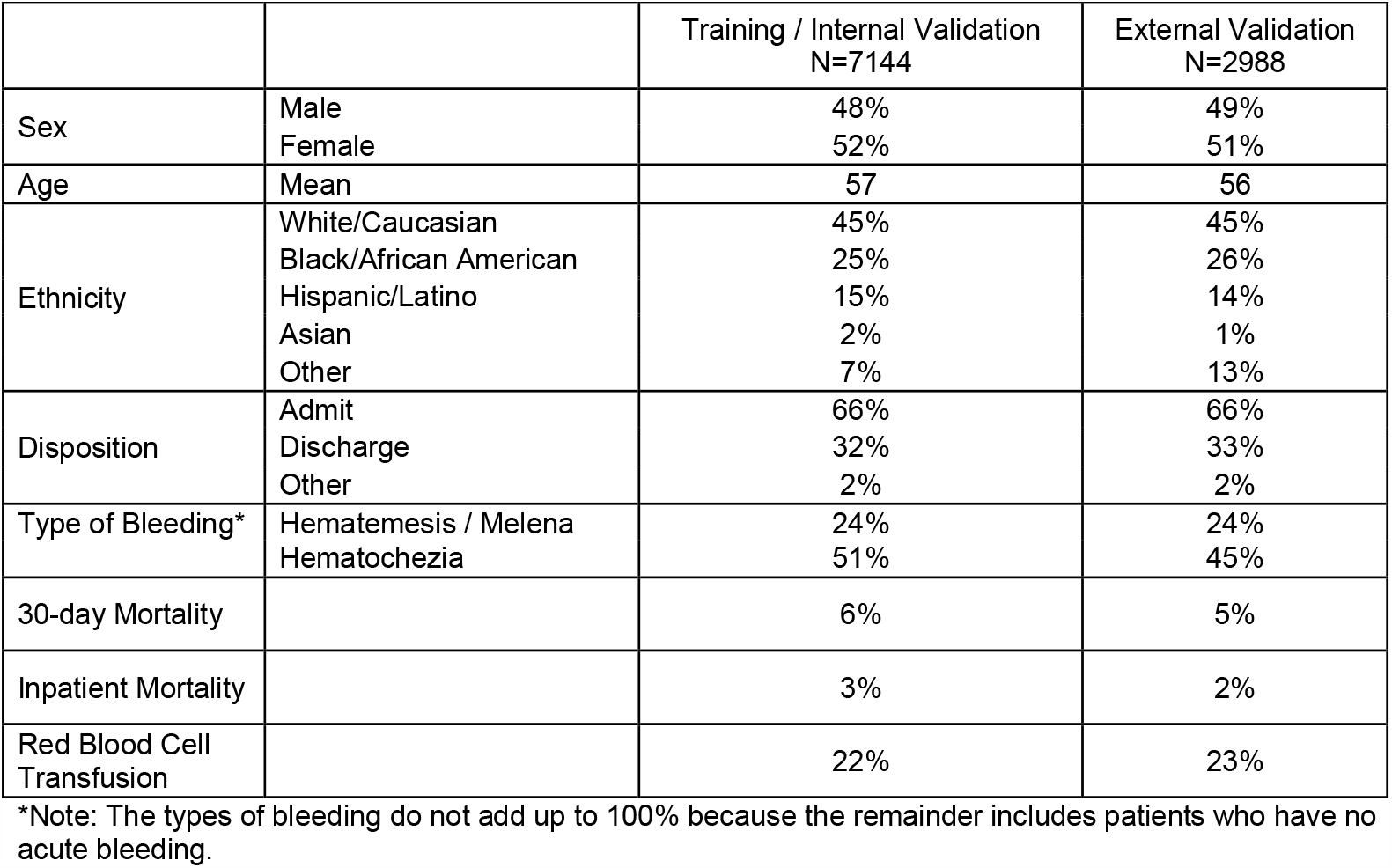
Baseline Characteristics for the Training and External Validation datasets

**Table 2:** Results of Phenotyping decision rules and NLP algorithm in Training and Validation Sets

|  |  |  |  |  |
| --- | --- | --- | --- | --- |
| Training (70%) N = 7144 |  |  |  |  |
| Internal Validation: Tenfold Cross-Validation |  |  |  |  |
| Phenotype and Performance Characteristics |  |  |  |  |
|  | Performance Algorithm (with 95% confidence interval) |  |  |  |
|  | SNOMED Codes<br>(reference)<br>Problem List<br>Encounter Diagnosis<br>Billing Diagnosis | Decision Rule 1:<br>All GI Triage<br>Terms + ROS<br>fields | Decision Rule 2:<br>GI Bleed-Specific<br>Triage terms +<br>ROS Fields | NLP only |
| PPV | 74%<br>(0.71 – 0.76) | 85%*<br>(0.84 – 0.87) | 91%*<br>(0.90 – 0.93) | 86%*<br>(0.82 – 0.90) |
| Sensitivity | 61%<br>(0.59 – 0.64) | 91%*<br>(0.90 – 0.93) | 88%*<br>(0.86 – 0.90) | 97%*<br>(0.96 – 0.98) |
| Specificity | 38%<br>(0.34 – 0.43) | 56%*<br>(0.53 – 0.59) | 77%*<br>(0.74 – 0.80) | 56%*<br>(0.48 – 0.64) |
| External Validation (30%) N = 2988 |  |  |  |  |
| Phenotype and Performance Characteristics |  |  |  |  |
|  | Performance Algorithm (with 95% confidence interval) |  |  |  |
|  | SNOMED Codes<br>(reference)<br>Problem List<br>Encounter Diagnosis<br>Billing Diagnosis | Decision Rule 1:<br>All GI Triage<br>Terms + ROS<br>fields | Decision Rule 2:<br>GI Bleed-Specific<br>Triage terms +<br>ROS Fields | NLP only |
| PPV | 69%<br>(0.67 – 0.71) | 78%*<br>(0.76 – 0.79) | 85%*<br>(0.84 – 0.87) | 80%*<br>(0.79 – 0.82) |
| Sensitivity | 59%<br>(0.57 – 0.61) | 90%*<br>(0.89 – 0.91) | 84%*<br>(0.82 – 0.85) | 98% *<br>(0.97 – 0.98) |
| Specificity | 45%<br>(0.42 – 0.48) | 47%<br>(0.43 – 0.50) | 71%*<br>(0.66 – 0.73) | 50%<br>(0.47 – 0.54) |

*P*<0.001 compared to SNOMED (baseline)

**Table 3:** Gold-Standard Strategy to Label Encounters

| Steps to label the Gold Standard: Acute Gastrointestinal Bleeding in the ED |
| --- |
| 1. Review of the ED Provider Note |
| 2. Any text that identifies acute gastrointestinal bleeding for |
| a. Hematemesis: e.g. “hematemesis”, “vomiting blood” |
| b. Melena: e.g. “dark stool”, “black stool”, “tarry stool”, “melena” |
| c. Hematochezia: e.g. “blood in toilet bowl”, “blood in stool”, “bright red blood in the stool” |
| 3. Patient report or physical exam findings were considered equally valid |
| 4. Exclude patients with other reasons for hematemesis – e.g. epistaxis |

### Training and Validation Datasets

The total number of encounters was temporally divided into training (70%, from 9/2014 to 7/2016) and validation (30%, 7/2016 to 5/2017) sets. Internal validation was performed with tenfold cross-validation across the training set for the NLP algorithm and the decision rules. External validation was performed directly using the held-out validation set. The primary metric for performance was the positive predictive value, and secondary metrics for performance the sensitivity and specificity. A high PPV indicates that a high proportion of the patients identified with acute GIB are true cases, but ideally would also have a high sensitivity to identify a high proportion of all true cases of acute GIB. There is no clear performance threshold for PPV, but PPV>75% has been considered acceptable and reported for EHR phenotypes.^13-18^

### Sensitivity Analysis by Bleeding Etiology and Prevalence

Pre-defined sensitivity analysis was performed to predict either hematemesis and/or melena or hematochezia, which are two clinically distinct symptom complexes that may indicate an upper gastrointestinal tract or lower gastrointestinal tract source. Separate NLP models were developed using the same methodology as for all acute GIB. Since PPV does depend on prevalence, a range of PPV was generated using prevalence from 1% to 100% and the sensitivity and specificity of each decision rule, and the lowest prevalence to maintain PPV > 75% calculated. Prevalence was defined with the numerator cases of acute GIB, and denominator total number of patients with any gastrointestinal problem as defined by the sensitive data mart. This approximates real life, where the patient population of interest is comprised of patients with a suspicion for an acute episode of GIB and not to detect a chronic condition in any patient presenting to the ED.

### Statistical Methodology

McNemar’s test was used to compare sensitivity, specificity, and PPV for external validation. The Wilcoxon signed-rank test, a nonparametric test for matched samples, was used for pairwise comparisons of sensitivity, specificity, and PPV for the tenfold internal cross-validation. PPV was considered the primary metric for performance with goal PPV >75%. We also predefined comparisons between the baseline (SNOMED) and each of the phenotyping approaches (decision rule 1, decision rule 2, and the NLP-based approach). We corrected using the Bonferroni correction and defined significance as *P*<0.01.

## Results

### Performance of Decision Rules

#### Internal Validation with Tenfold Cross-Validation

Decision rules 1, 2, and the NLP-based tool had significantly better PPVs than SNOMED codes in identifying patients with acute bleeding. The NLP-based tool had the best sensitivity (0.97 CI 0.96-0.98) and was significantly better than SNOMED (0.61 CI 0.59-0.64, *P*<0.001). Decision Rule 1 (0.91 CI 0.90-0.93) and Decision Rule 2 (0.88 CI 0.86-0.90) also have significantly higher sensitivity than SNOMED. Decision Rule 2 had the best specificity (0.77 CI 0.74-0.80), which was significantly better than SNOMED (0.38 CI 0.34-0.43, *P*<0.001). Both Decision Rule 1 (0.56 CI 0.53-0.59) and the NLP-based tool (0.56 CI 0.48-0.64) also have significantly higher specificity than SNOMED (*P*<0.001).

#### External Validation

The PPV of decision rules (78% for decision rule 1, 85% for decision rule 2) and NLP (80%) were significantly increased compared to SNOMED (69%; *P*<0.001). The sensitivity of decision rules (90% for decision rule 1, 84% for decision rule 2) are significantly increased compared to SNOMED codes (59%;*P*<0.001). For specificity, SNOMED codes (46%) is significantly worse than decision rule 2 (71%;*P*<0.001) but similar to decision rule 1 (47%;*P*=0.87) and the NLP tool (50%;*P*=0.84).

### Sensitivity Analysis and Prevalence Range

#### Sensitivity Analysis based on Type of Bleeding

On external validation for hematemesis and/or melena alone the decision rules performed similarly to SNOMED codes and the NLP tool had the best PPV, which reflected a very high specificity but lower sensitivity than SNOMED codes and either of the decision rules. (Table 4)

**Table 4:** Sensitivity Analysis of Hematemesis and/or Melena and Hematochezia in the External Validation Group

| Hematemesis and/or Melena Validation (30%) N = 2988 |  |  |  |  |
| --- | --- | --- | --- | --- |
| Phenotype and Performance Characteristics |  |  |  |  |
|  | Performance Algorithm (with 95% confidence interval) |  |  |  |
|  | SNOMED Codes (reference) | Decision Rule 1: All GI Triage Terms + ROS fields | Decision Rule 2: GI Bleed-Specific Triage terms + ROS Fields | NLP only |
| Sensitivity | 71%<br>(0.67 – 0.74) | 77%<br>(0.73 – 0.80) | 68%<br>(0.64 – 0.71) | 27%<br>(0.24 – 0.31) |
| Specificity | 46%<br>(0.44 – 0.48) | 22%<br>(0.20 – 0.23) | 35%<br>(0.33 – 0.37) | 99%<br>(0.98 – 0.99) |
| PPV | 28%<br>(0.26 – 0.31) | 23%<br>(0.21 – 0.25) | 24%<br>(0.22 – 0.26) | 86%*<br>(0.80 – 0.90) |
| Hematochezia Validation (30%) N = 2988 |  |  |  |  |
| Phenotype and Performance Characteristics |  |  |  |  |
|  | Performance Algorithm (with 95% confidence interval) |  |  |  |
|  | SNOMED Codes (reference) | Decision Rule 1: All GI Triage Terms + ROS fields | Decision Rule 2: GI Bleed-Specific Triage terms + ROS Fields | NLP only |
| Sensitivity | 55%<br>(0.52 – 0.57) | 96%<br>(0.95 – 0.97) | 91%<br>(0.90 – 0.93) | 79%<br>(0.77 – 0.81) |
| Specificity | 40%<br>(0.37 – 0.42) | 37%<br>(0.34 – 0.39) | 54%<br>(0.52 – 0.56) | 89%<br>(0.88 – 0.91) |
| PPV | 42%<br>(0.39 – 0.44) | 55%*<br>(0.53 – 0.57) | 61%*<br>(0.59 – 0.63) | 85%*<br>(0.83 – 0.87) |

*P*<0.001 compared to SNOMED (baseline)

For hematochezia alone, the NLP tool similarly had the best PPV but lower sensitivity than SNOMED codes and either of the decision rules. The decision rules outperformed SNOMED for PPV (DR1 55% and DR2 61% versus SNOMED 42%, p<0.001), but had similar specificity. (Table 4)

#### Positive Predictive Value and Prevalence

For the optimal decision rule in the external validation set, the PPV was calculated with a prevalence of 67%. This prevalence reflects an enriched population of patients with any indication of GIB during their hospital stay. When tested across the range of prevalence from 1% to 100%, the PPV was >75% with a prevalence as low as 50%.

## Discussion

This is the first study to develop an EHR phenotype for acute gastrointestinal bleeding that could be used in the emergency room to deploy risk scores to inform determine level of care and inform clinical management decisions. (Figure 3) We found that the decision rule that combined bleed-specific terms at initial triage by a nurse and bleed-specific terms in the emergency department provider’s review of systems had the highest PPV (85%) on external validation with a sensitivity of 84% to identify patients presenting with acute GIB in the emergency department. This decision rule can be used to trigger alerts that can help triage patients into high and low risk.

**Figure 3:**
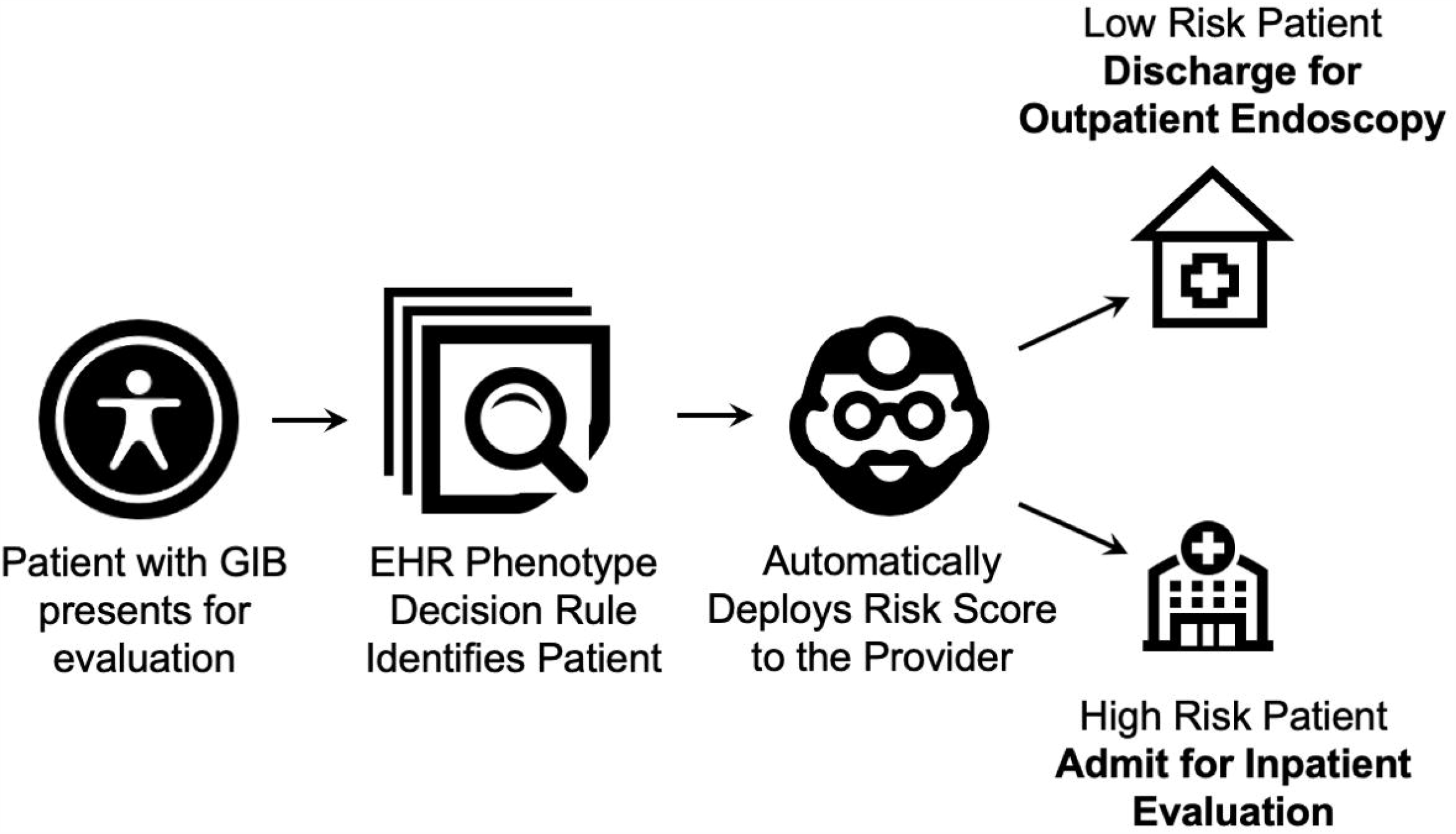
Schematic showing how the electronic health record (EHR) phenotyping rule fits into the provider workflow to find patients with gastrointestinal bleeding and deploy risk scores to assist decision making.

We chose the decision rule because in practice we want to deploy a risk prediction tool for patients who actually have acute GIB, which means we want a low false positive rate (high specificity and positive predictive value). Sensitivity should also be as high as possible to ensure that most patients with acute GIB are detected. A positive predictive value of >75% has been reported as “high,” and the PPV for the highest performing decision rule was 91% on internal validation and 85% on external validation.^13-18^

Traditionally no method exists to identify patients with gastrointestinal bleeding other than using diagnostic codes (ICD-9, ICD-10) from the billing diagnosis list. By definition, these diagnostic codes are useless for detecting patients presenting with acute GIB at the point of care because they generally are not entered into the EHR until much later in the hospital course, well after initial identification and risk assessment is needed. Additionally, ICD codes may not detect all patients who have bleeding, as it may be reported but not coded in their billing diagnosis. SNOMED may be better than diagnostic codes alone—it is a terminology that includes both billing codes and other related text.^19,20^ Our decision rule outperforms the baseline SNOMED methodology even when sampled over multiple areas in the EHR, including the billing diagnosis list. Interestingly, introduction of NLP over the entire text of the emergency department provider notes has a much higher sensitivity but even in conjunction with the decision rules does not have high specificity. Furthermore, the availability of the entire text is variable, since providers may choose to complete documentation up to 24 hours after the actual visit.

The decision for need, timing, and type of endoscopy may be based on presumptive diagnosis (e.g. upper endoscopy versus colonoscopy), but the initial management is similar for both UGIB and LGIB. The level of care, resuscitation, and optimization of other active co-morbidities are relevant prior to any endoscopic evaluation, EGD and colonoscopy are both recommended for most patients admitted with GIB. The sensitivity analysis suggested that neither decision rule performs well to identify symptoms associated with UGIB (hematemesis or melena) or LGIB (hematochezia). The NLP tool had a higher PPV both UGIB and LGIB, but is not suited for prospective deployment in real time to identify patients with GIB. The sensitivity of NLP for upper GIB was incredibly low (27%), reflecting the fact that the tool would incorrectly classify more than half of patients with the symptoms as negative. The NLP tool performed better for lower GIB, and could be explored further in a separate study. The role of novel tools such as NLP continues to advance, and we are hopeful that new techniques and tools will lead to improved performance.

### Strengths

This study compares the performance of existing automated methods (SNOMED) across multiple time points in the EHR in identifying acute GIB at admission. This excludes GIB after admission while already hospitalized for another condition, and provides the best data possible given the constraints of using only structured datafields to develop risk stratification scores for patients who present with acute GIB to the emergency department.

### Limitations

We did not review patients who did not have the specific SNOMED code for GIB, did not have a GI-related triage problem, and did not have a positive review of systems for GIB. We believe it is reasonable to assume that these patients likely present with another primary issue and without any clinically significant GIB. Risk stratification scores for acute gastrointestinal bleeding are typically used for patients who present with GIB as the chief and acute complaint, and clinical decisions, such as admission or hospital-based interventions, need to be made early after presentation.

This phenotype was developed for a specific center with a local workflow, including the availability of a triage nurse with structured datafields for triage diagnosis. Patients outside this cohort (with negative SNOMED, none of the triage terms, and no ROS positivity) were not reviewed, which limits its applicability to all-comers in the emergency department. However, we believe patients without any GI symptoms or signs at triage or during emergency department evaluation and without any evidence of GIB on diagnostic codes or the other elements of SNOMED are very unlikely to have presented with clinically significant acute GIB. Changes in coding (e.g. from ICD-9 to ICD-10) and temporal shifts in treatment options, patient epidemiology, hospital utilization, and risk shifts can all decrease the performance of these phenotypes in identifying patients of interest.

### Future Directions

These results provide a robust method of identifying patients with acute GIB to develop an EHR-based model for risk prognostication. By mapping out the points at which data is generated from the process model, this guides the eventual deployment and implementation of a risk stratification prognostic algorithm for clinical decision making in real time.

## Data Availability

All patient data is stored on encrypted and password-secured storage and is not publicly available to preserve patient privacy.

## Acknowledgements

We express our gratefulness to J. Kenneth Tay (Stanford University) for advice on statistical methodology.

## Notes

Funding: DS is supported by NIH grant T32 DK007017.

### Competing Interest Statement

The authors have declared no competing interest.

### Funding Statement

DS is supported by NIH grant T32 DK007017.

### Author Declarations

Institutional Review Board of Yale University, Human Investigation Committee HIC # 1408014519

